# Motion-robust high-resolution 3D-GRASE pseudo-continuous ASL perfusion imaging at 3T

**DOI:** 10.64898/2026.10.23.26366363

**Authors:** Minhao Hu, Frederik J. Lange, Peter Jezzard, Mark Chiew, Thomas W. Okell

## Abstract

**Purpose:** High resolution arterial spin labeling (ASL)-based perfusion imaging is desirable for many applications but suffers from low SNR and motion sensitivity due to the necessity for highly segmented readouts. Acquisition and reconstruction strategies were optimized for motion-robust high-resolution pseudo-continuous arterial spin labeling (pCASL) of the brain at 3T using segmented 3D Gradient and Spin Echo (3D-GRASE) at 2.0-mm isotropic resolution.

**Methods:** A high-resolution 3D-GRASE pCASL sequence was implemented at 3T and evaluated with a focus on the interaction between background suppression strength and motion-corrected reconstruction, together with the fat suppression and control-tag subtraction strategies that strong suppression requires. Motion correction was performed using a novel motion-compensated (alignedSENSE-based) reconstruction that simultaneously estimates the motion and image, accounting for contrast changes between tag and control data. Eight healthy volunteers were scanned under motion-free and cued motion conditions. Image quality was assessed against motion-free reference images using peak signal-to-noise ratio (PSNR) and a structural similarity index measure (SSIM).

**Results:** The optimized acquisition-reconstruction pipeline enabled robust 2.0-mm isotropic pCASL at 3T and showed improved resistance to motion-related degradation. Strong BGS provided near-complete suppression of static tissue, and water excitation reduced lipid contamination and improved coil sensitivity estimation. The regularized alignedSENSE motion correction further improved perfusion-image quality, increasing SSIM by 18.7% under Strong BGS.

**Conclusion:** Robust high-resolution pCASL at 3T can be achieved through combined optimization of acquisition and reconstruction. The combination of Strong BGS, water excitation, reference-superimposed processing, and regularized motion correction mitigates motion-related degradation and yields higher-quality perfusion images for high-resolution ASL.

## INTRODUCTION

Cerebral perfusion, the delivery of blood to brain tissue, is a fundamental physiological parameter that is altered in a wide range of neurological and cerebrovascular disorders, including stroke^1^, dementia^2^, brain tumors^3^, and neurodegenerative disease^4^. Perfusion imaging has traditionally relied on exogenous contrast agents, such as dynamic susceptibility contrast MRI^5^ or CT perfusion^6^, which carry risks related to contrast administration^7^ and are less suitable for repeated or longitudinal studies. Higher spatial resolution is desirable in perfusion imaging as it may improve the detection of focal abnormalities, laminar perfusion differences, and small-vessel pathology that can be obscured at conventional resolutions^8^.

Arterial spin labeling (ASL) MRI provides a fully non-invasive alternative by using magnetically labeled arterial water as an endogenous tracer^9,10^. Because no exogenous contrast agent is required, ASL is well suited to repeated measurements, longitudinal studies^11^, and vulnerable patient groups such as pediatric^12^ or renally impaired populations^13^. Pseudo-continuous ASL (pCASL)^14^ has become the recommended labeling strategy due to its high labeling efficiency and favorable SNR^15^. At conventional spatial resolutions (typically 3–4 mm isotropic), ASL has been successfully applied in a broad range of clinical and neuroscientific settings, including assessment of cerebrovascular reactivity^16^, tumor characterization^17^, and resting-state brain function^18^.

Extending ASL to higher spatial resolution remains challenging. The perfusion signal is intrinsically small, typically on the order of 1-2% of the tissue signal^15^, and this low SNR becomes more limiting as voxel size decreases. High-resolution imaging also generally requires longer readouts or more highly segmented acquisitions, which increase sensitivity to physiological fluctuation and head motion. In particular, segmented 3D readouts such as 3D Gradient and Spin Echo (3D-GRASE)^19–21^ are attractive for high-resolution ASL because of their acquisition efficiency and compatibility with background suppression (BGS), but they require multiple excitations to fill k-space, making the final perfusion image vulnerable to inter-shot motion^22^. As a result, high-resolution ASL at 3T is constrained not by a single factor, but by the combined effects of low SNR, physiological noise, and motion sensitivity.

Previous work has advanced high-resolution ASL through segmented 3D readouts, optimized BGS, acceleration and regularized reconstruction. At 3T, segmented 3D acquisitions achieved 2 × 2 × 4 mm^3^ resolution^23^, while controlled aliasing in parallel imaging (CAIPI)^24^ accelerated 3D-GRASE with z-deblurring enabled 2-mm isotropic whole brain imaging in about 5 min^8^. Spatio-temporal total generalized variation denoising^25^ further improved low-SNR ASL, and optimized 7T pCASL with dynamic compressed sensing has reached 0.8-mm isotropic resolution^26^. These studies primarily addressed SNR and acceleration; head motion was generally assumed to be small or handled by conventional retrospective registration. However, segmented 3D-GRASE is inherently sensitive to inter-shot motion. Huber et al. jointly estimates motion and distortion^27^, and we previously introduced self-navigated inter-shot correction for segmented 3D-GRASE ASL with CAIPI sampling^28^. alignedSENSE^29^ offers a complementary approach by jointly estimating the image and rigid-body motion from multicoil k-space and has been extended to 3D multi-shot EPI^30,31^. These formulations, however, assume common contrast across shots and do not account for the systematic intensity difference between tag and control data in ASL or how BGS strength affects motion estimation.

Background suppression is central to this problem. Strong background suppression is attractive for high-resolution ASL because it lowers the physiological noise arising from incompletely subtracted static tissue, and by reducing the residual CSF signal^32^, limits the amount of background signal that head motion can convert into subtraction artifacts. This suppression, however, removes most of the static-tissue contrast on which image-based registration depends, so the degree of suppression that is most favorable for the perfusion measurement is also the most challenging for conventional motion correction. More moderate suppression retains anatomical contrast that aids motion estimation but leaves more residual static signal that can be transformed into artifacts when motion occurs. This trade-off is not resolved by simply extending our previous self-navigation approach^28^, because at 2.0 mm isotropic resolution each shot is too heavily undersampled to provide the self-navigating information that method relies on. The interaction between background suppression strength and motion-correction therefore remains undetermined.

In this work, we address this question for high-resolution 3D-GRASE pCASL at 3T. We propose alignedSENSE-PR, which adds perfusion regularization (PR) to alignedSENSE and estimates inter-shot motion directly from the multicoil k-space data through the forward model. The PR term accounts for the systematic contrast difference between tag and control images, helping to maintain robust tag-control alignment even when strong background suppression removes most image-domain tissue contrast. This allows strong background suppression to be used to its full advantage for high-resolution ASL. Adopting strong suppression in turn places specific demands on the rest of the pipeline: residual lipid becomes proportionally more conspicuous, requiring effective fat suppression through water excitation, and the suppressed signal can approach or span zero, so that magnitude subtraction becomes biased and a sign-preserving subtraction strategy is needed. We therefore evaluate background suppression strength, fat suppression, control-tag subtraction method, and motion-corrected reconstruction together at 2.0 mm isotropic resolution and identify the combination that yields the most robust perfusion image quality under conditions of head motion.

## METHODS

### Sequence Design

An overview of the sequence design is shown in **Figure 1a**. A 3D-GRASE^21,33^ pCASL sequence previously developed in our group was further modified and implemented on a 3T scanner. The labeling parameters were labeling duration (LD) = 1800 ms, and post-label delay (PLD) = 1800 ms.

**Figure 1.**
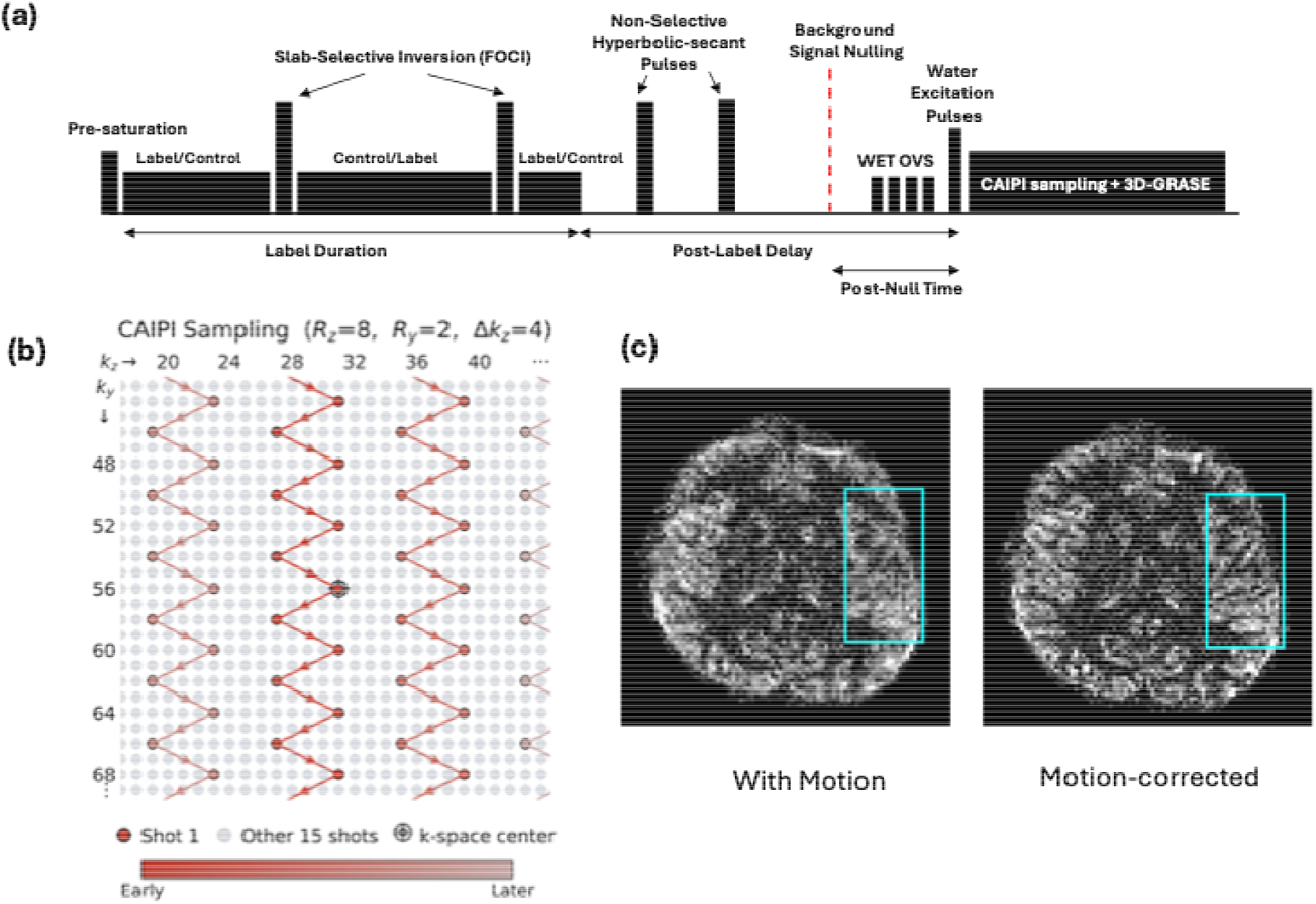
Overview of the optimized high-resolution pCASL sequence and motion correction framework. (a) Pulse sequence diagram. A pCASL labeling scheme is followed by a post-label delay. Four inversion pulses are applied for background suppression (BGS): the pulses interleaved with the pCASL pulse train are slab-selective frequency-offset corrected inversion (FOCI) pulses, while the remaining pulses are non-selective hyperbolic-secant pulses. The post-null time (PNT) is defined as the interval between the targeted null time (red dashed line) and the excitation. Water excitation and outer volume suppression (OVS) pulses precede the segmented 3D-GRASE readout with CAIPI sampling. (b) Illustration of the time-dependent CAIPI k-space sampling pattern (R**y = 8**,R**z =** 2, Δ**z** = 4). k-space is divided into 16 segments; one representative shot (red) is shown alongside the remaining 15 shots (grey), demonstrating the interleaved shifting of the ky– kz sampling trajectory across segments. (c) Representative perfusion-weighted images from a single subject, illustrating motion-induced artifacts (left) and the improvement achieved by the motion-corrected reconstruction method (right). The boxes highlight regions most affected by motion.

Background suppression used a four-pulse (4-inv BGS)^34^ scheme applied after pre-saturation and before excitation. The inversion pulses interleaved with the pCASL pulse train were slab-selective frequency-offset-corrected inversion (FOCI)^35^ pulses to achieve spatially uniform inversion proximal to the labeling plane with sharp edges, while the remaining inversion pulses were non-selective hyperbolic-secant pulses. After each inversion pulse, the labeling state was inverted accordingly. The degree of background suppression was controlled by the post-null time (PNT), defined as the interval between the theoretical signal nulling point and the excitation pulse, which determines the level of residual longitudinal magnetization at the time of readout. Two 4-inv BGS timing schemes were evaluated. In the “**Moderate BGS**” condition, tissue signals were nulled 100 ms before excitation (PNT = 100 ms), allowing partial longitudinal recovery and resulting in residual background signal. In the “**Strong BGS**” condition, tissue signals were nulled at the time of excitation (PNT = 0 ms), maximizing background suppression. The inversion pulse timings were computed using the coefficients of the quadratic polynomial equations described by Maleki et al.^34^.

A 1-2-1 binomial water excitation pulse^36^ was used for fat suppression. Four outer volume suppression (OVS) pulses were applied prior to the readout to reduce aliasing from outside the imaging slab^21^.

A segmented 3D-GRASE readout^21,33^ was employed with a time-dependent CAIPI 2×8(4) sampling pattern^28,37^ (**Figure 1b**) with in-plane acceleration *R*_*v*_ = 8, through-plane acceleration *R*_*z*_ = 2, and *k*_*z*_ sampling steps Δ _*z*_= 4. k-space was divided into 16 segments, with turbo factor = 8 and EPI factor = 57. The sampling indices begin at the k-space center and step outward, with the ky–kz trajectory shifted between segments so that each shot samples the full k-space volume rather than a localized region. This uniform coverage maximizes the information each shot provides for estimating and correcting its own motion, while also distributing the undersampling artifacts incoherently across k-space. The time-dependent CAIPI sampling strategy has been successfully adopted in several recent studies for both single-shot and segmented 3D acquisitions^28,37,38^. In addition, the complete k-space coverage across shots allows all segments to be combined to form a fully sampled dataset for robust coil sensitivity calibration. Readout parameters were refocusing flip angle = 120°, TR = 4 s, TE = 46.5 ms, bandwidth = 1994 Hz/px, matrix size = 114 × 114 × 64, yielding 2.0 mm isotropic resolution.

Tag and control encoding was interleaved shot-by-shot before advancing the sampling mask, so that corresponding tag and control segments were acquired in close temporal proximity, minimizing motion-related misalignment^15^.

### Background Suppression Simulation

Residual longitudinal magnetization through the BGS pulse train was simulated as a function of tissue *T*_*1*_ to characterize the BGS configurations. The simulation modelled *T*_*1*_ recovery from an ideal pre-saturation pulse *(M*_*z*_ = 0), followed by the action of the inversion pulses and subsequent recovery to the excitation time point. Inversion pulses were modelled with an inversion efficiency of 95%. For comparison, a double-inversion scheme with PNT = 100 ms (2-inv BGS) was simulated using the Günther et al.^21^ formulation with *T*_*l,opt*_ = 700 *ms*. The corresponding four-inversion simulations used the Maleki et al.^34^ polynomial timing model. The three simulated configurations: 2-inv BGS, Moderate BGS, and Strong BGS matched the three conditions implemented in vivo. Simulations used LD =1800 ms and PLD = 1800 ms, with residual *M*_*z*_*/M*_0_ evaluated at excitation. Approximate tissue *T*_*1*_ ranges at 3T were used to interpret the residual-signal curves: lipid, 200-400 ms; white matter, 800-1100 ms; grey matter, 1200-1600 ms; and CSF, 3500-4000 ms.

### Perfusion-Regularized Motion-Corrected Reconstruction

As tag and control data were acquired in an interleaved shot-by-shot fashion, a complete tag or control volume used for reconstruction extended over 2 min. In contrast, the acquisition window of each individual shot was relatively short. Motion was therefore assumed to be dominated by inter-shot displacement, and only rigid-body motion between shots was explicitly modelled, while intra-shot motion was ignored. At this spatial resolution a navigator image cannot be reconstructed from each individual shot^28^, so motion is instead estimated jointly with the image as part of the reconstruction, using alignedSENSE-PR, a perfusion-regularized variant of alignedSENSE^29^.

A single mean image *x* and perfusion image Δ*x* were shared across all three tag-control repeats, with rigid-body motion parameterized per repeat and shot by transformation matrices *T*_*p,r,i*_ *(ASL state p∈ {t, c}*, repeat *r∈*[*t*, c], *shot i∈*[1, 16]). The tag and control images are modelled as *x — Δx* and *x + Δ x*, respectively. Sharing one perfusion estimate across repeats maximizes the supporting data and yields globally consistent, robust motion estimation. The reconstruction was framed as the following optimization:

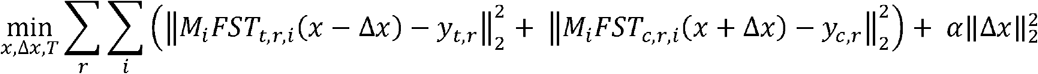

where *M*_*i*_ is the undersampling operator for the *i*-th sampling mask, *F* the Fourier transform, *S* the coil sensitivity maps, *y*_*t,r*_ and *y*_*c,r*_ the k-space measurements for tag and control, respectively, and *α* the regularization weight applied to the perfusion image.

The L2 penalty on *Δx* has a specific role beyond noise suppression. When inter-shot motion is correctly estimated, static tissue is consistent across shots and between tag and control, so the only systematic control–tag difference is the true perfusion signal; any residual misalignment instead introduces spurious static-tissue differences that inflate *Δ x*. Penalizing ∥*Δx*∥_2_ therefore drives the joint optimization toward motion parameters that minimize this artifactual static-tissue subtraction, stabilizing motion estimation precisely in the Strong BGS regime where the large control–tag contrast change would otherwise corrupt it.

The problem was solved iteratively, alternating between three inner image update steps and one motion estimation step per outer iteration, with a stopping tolerance of 10^−6^ and a maximum of 50 outer iterations. The regularization weight *α* = 1 was empirically selected based on reconstruction quality (see Supporting Information **Figure S1**) and was used for all subsequent experiments.

The perfusion image Δ*x* estimated in this step plays no role in the final perfusion measurement and its only function is to stabilize the motion estimation. Any bias introduced by the L2 penalty therefore affects the estimated motion parameters alone, not the reported perfusion signal, which was formed in a separate step from the estimated motion using a reference-superimposed (RS) reconstruction^39^.

Because alignedSENSE-PR corrects both inter-volume and inter-shot motion of the tag and control data, its output can be brought into alignment with the non-suppressed equilibrium-magnetization reference (*M*_*0*_), allowing the *M*_*0*_ k-space to be superimposed onto each volume. Adding the *M*_*0*_ signal, which shares the phase of the underlying tissue, lifts the tag and control signals away from the null point before the magnitude is taken, so that magnitude subtraction no longer suffers the sign ambiguity and noise rectification that can arise under Strong BGS when the residual longitudinal magnetization approaches or spans zero.

Concretely, the k-space data from the two *M*_*0*_ repeats were averaged and then added to each aligned tag and control dataset; each dataset was reconstructed and converted to magnitude, and the perfusion-weighted image was formed by magnitude subtraction. All reconstructions were mapped to a common motion-free reference space for metric computation as described in the Quantitative Evaluation section, and the *M*_*0*_ reference was acquired in this same space.

No denoising was applied to any result reported in the main text. However, an optional locally low-rank (LLR) constraint, which exploits the redundancy of the perfusion component across repeats, is described in Supporting Information **Text S1**.

Reconstruction was performed on a workstation with 4 AMD EPYC 9274F CPUs (24 cores) and 128 GB RAM, requiring approximately 20 min per reconstruction.

## Data Acquisition

Eight healthy volunteers (1 female; age: 29.6 ± 3.2 years) were scanned on a 3T Prisma scanner (Siemens Healthineers) with a 32-channel head coil. All imaging was conducted under a technical development protocol agreed by local ethics and institutional committees.

Each subject underwent four ASL acquisitions after the *M*_0_ scan in a fixed order: Strong BGS without instructed motion, Moderate BGS without instructed motion, Strong BGS with cued motion and Moderate BGS with cued motion. For the first two acquisitions, subjects were instructed to remain stationary throughout and these data served as nominal motion-free references. For the last two acquisitions, subjects performed cued moderate head motion^28,40^, were used for motion correction evaluation. Each acquisition comprised three tag-control pairs and lasted 6 min 29 s.

An *M*_0_ reference was acquired at the start of each session with labeling and background suppression disabled, using the same 3D-GRASE readout, CAIPI sampling, and spatial resolution as the ASL acquisitions (TR/TE = 5500/46.54 ms). Two repeats were acquired (total acquisition time = 2 min 56 s) and averaged to form the *M*_0_ reference. This *M*_0_ reference was shared across the Strong BGS, Moderate BGS, motion-free, and motion-corrupted acquisitions, and provided the reference for the reference-superimposed reconstruction described above.

In each session, a T1-weighted anatomical image was acquired for registration and tissue segmentation (resolution: 1.7 mm isotropic; FOV = 220 × 200 × 218 mm; TR/TE/TI = 1900/3.71/904 ms; flip angle = 8°; acquisition time = 3 min 40 s).

### Quantitative Evaluation

To allow image-similarity metrics to be compared fairly across reconstruction methods, the output of every method was brought into a common reference space before evaluation. This reference space was defined by the temporal average of the tag and control volumes from the motion-free reference acquisition. For each method, each reconstructed tag/control volume was registered to this motion-free reference with FSL registration tool FLIRT^41^ to obtain a residual rigid-body transformation. This residual transformation was then composed with the intra-scan motion parameters estimated by that method, and a final reconstruction was run with the combined motion, mapping every method’s output into the identical reference space whilst avoiding image-space interpolation effects.

Brain masks were extracted by using FSL brain-extraction tool BET^42^ from T1-weighted images and then registered to the reference space. Image quality was assessed by computing two metrics within the brain mask against the motion-free reference: the Structural Similarity Index Measure (SSIM) and the Peak Signal-to-Noise Ratio (PSNR). Both metrics were computed on the perfusion-weighted image.

Statistical significance of differences between reconstruction methods was assessed using paired-sample t-tests, with significance levels set at p < 0.05 (*), p < 0.01 (**), and p < 0.001 (***).

### Comparison Conditions

Having established Strong BGS as the target operating regime, we evaluated a set of baseline and ablation conditions to test whether each accompanying choice is required in that regime and to isolate the contribution of the proposed reconstruction.

For background suppression, 2-inv BGS was included as a reference for Moderate and Strong BGS, consistent with prior ASL implementations^21,43^.

Fat suppression compared the vendor-provided fat saturation with 1-2-1 water excitation. Additional binomial and no-fat-suppression conditions were evaluated at 3.6-mm isotropic resolution (**Figure S2**).

For reconstruction ablation, alignedSENSE-PR was compared with alignedSENSE-IS (inter-shot correction only) and alignedSENSE-MF (inter-shot correction was followed by mcFLIRT^41^ image-based inter-volume registration). Because RS requires tag, control, and *M*_*0*_ k-space in a common frame, the ablation used complex subtraction, which is applicable to all variants (**Figure S3**).

Complex subtraction was also evaluated as an alternative to RS with and without LLR denoising (**Figures S4 to S6**).

## RESULTS

### Background Suppression

**Figure 2a** shows the simulated residual longitudinal magnetization *(M*_*z*_*/M*_0_) as a function of tissue T1 for the three BGS configurations evaluated in vivo. At the matched PNT of 100 ms, 2-inv BGS provided only limited suppression, leaving 8.9% residual CSF signal, whereas Moderate BGS reduced the residual CSF signal to 2.6%. Strong BGS reduced it further to - 0.2% and achieved near-complete suppression across tissue types.

**Figure 2.**
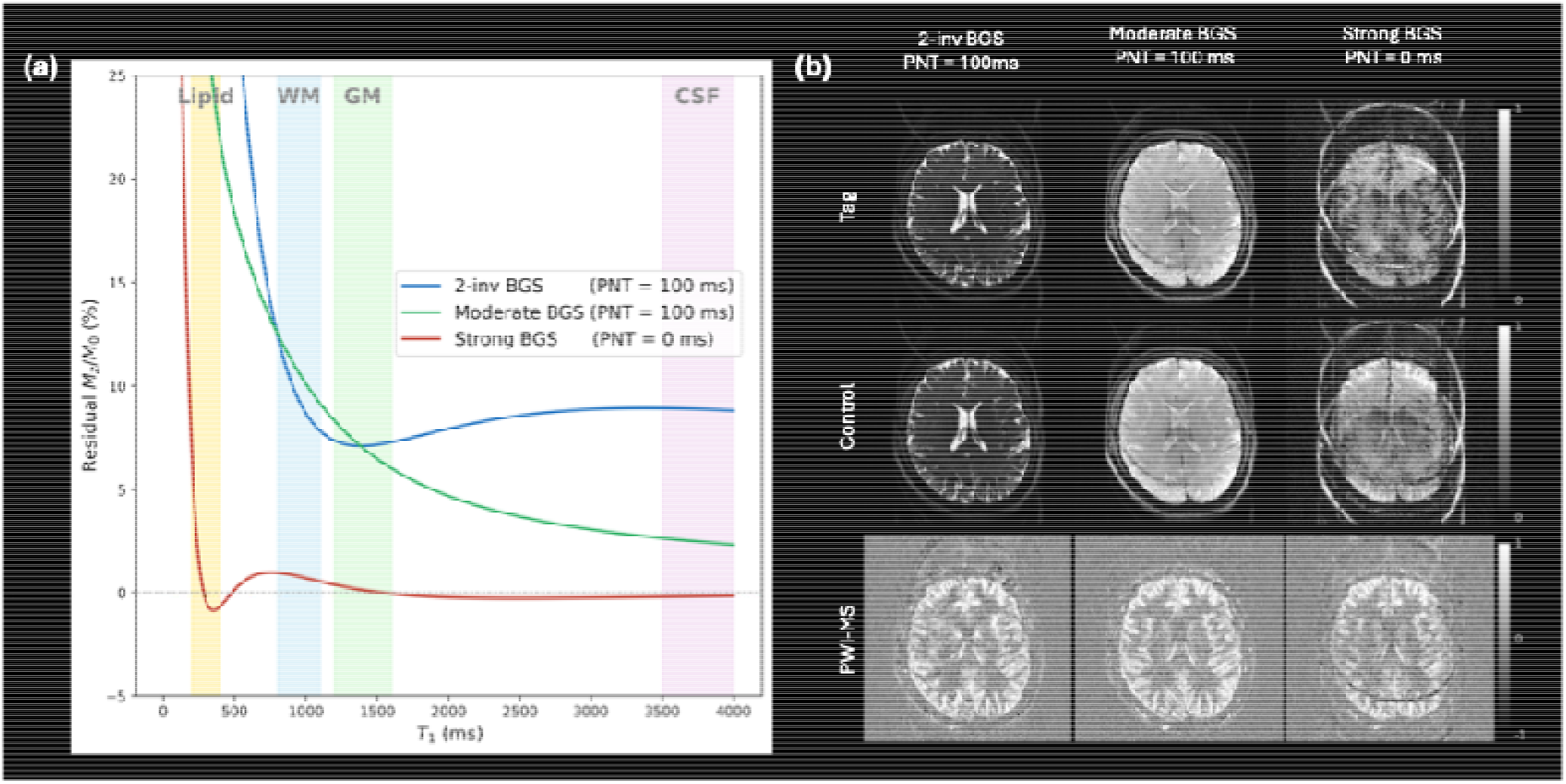
Background suppression simulation and in vivo images. (a) Simulated residual longitudinal magnetization **(M**_**z**_**/M**_**0**_ as a function of tissue **T**_1_ for 2-inv BGS (blue), Moderate BGS (green) and Strong BGS (red). Shaded regions indicate the approximate **T**_1_ ranges for lipid, white matter (WM), grey matter (GM), and CSF at 3T. (b) Representative tag, control, and perfusion-weighted images from a single subject acquired under these three configurations in vivo. Note that these images were obtained using conventional fat saturation and magnitude subtraction, isolating the effect of BGS scheme alone. Water excitation and subtraction strategy are introduced and evaluated independently in subsequent sections.

The representative in vivo images in **Figure 2b** (normalized individually for visualization) are consistent with the simulation. Compared to 2-inv BGS, Moderate BGS substantially reduced CSF signal in the ventricles in both tag and control images, lowering physiological noise in the perfusion-weighted image (PWI). Strong BGS further reduced the static background and increased the intensity difference between tag and control images. As 2-inv BGS provided substantially inferior suppression, it was included only as a visual reference and was not carried forward into the quantitative motion correction analysis.

Strong BGS leads to changes in image contrast and motion sensitivity. First, once the tissue background is strongly suppressed, residual lipid signal becomes proportionally more conspicuous and appears as a ring-like structure at the periphery, motivating the fat suppression comparison in the following section. Second, the suppressed tissue also operates in a low-signal regime in which the residual longitudinal magnetization can approach zero or reverse sign, motivating the subtraction strategy comparison in the following sections. Third, the strong reduction of static-tissue contrast that makes the subtraction cleaner also removes much of the anatomical information on which conventional image-based motion correction relies, which is the central tension addressed by the proposed reconstruction.

### Fat Suppression: Fat Saturation vs Water Excitation

**Figure 3a** compares reconstructed images obtained with conventional fat saturation and with water excitation. Under fat saturation, residual chemically-shifted lipid signal was incompletely suppressed and appeared as focal hyperintensities in the tag and control images (yellow arrows), predominantly in regions adjacent to subcutaneous fat. These signals propagated into the PWI as structured artifacts (yellow box) that were not removed by magnitude subtraction. In contrast, water excitation effectively eliminated lipid signal, resulting in cleaner tag and control images and improved spatial homogeneity in the PWI.

**Figure 3.**
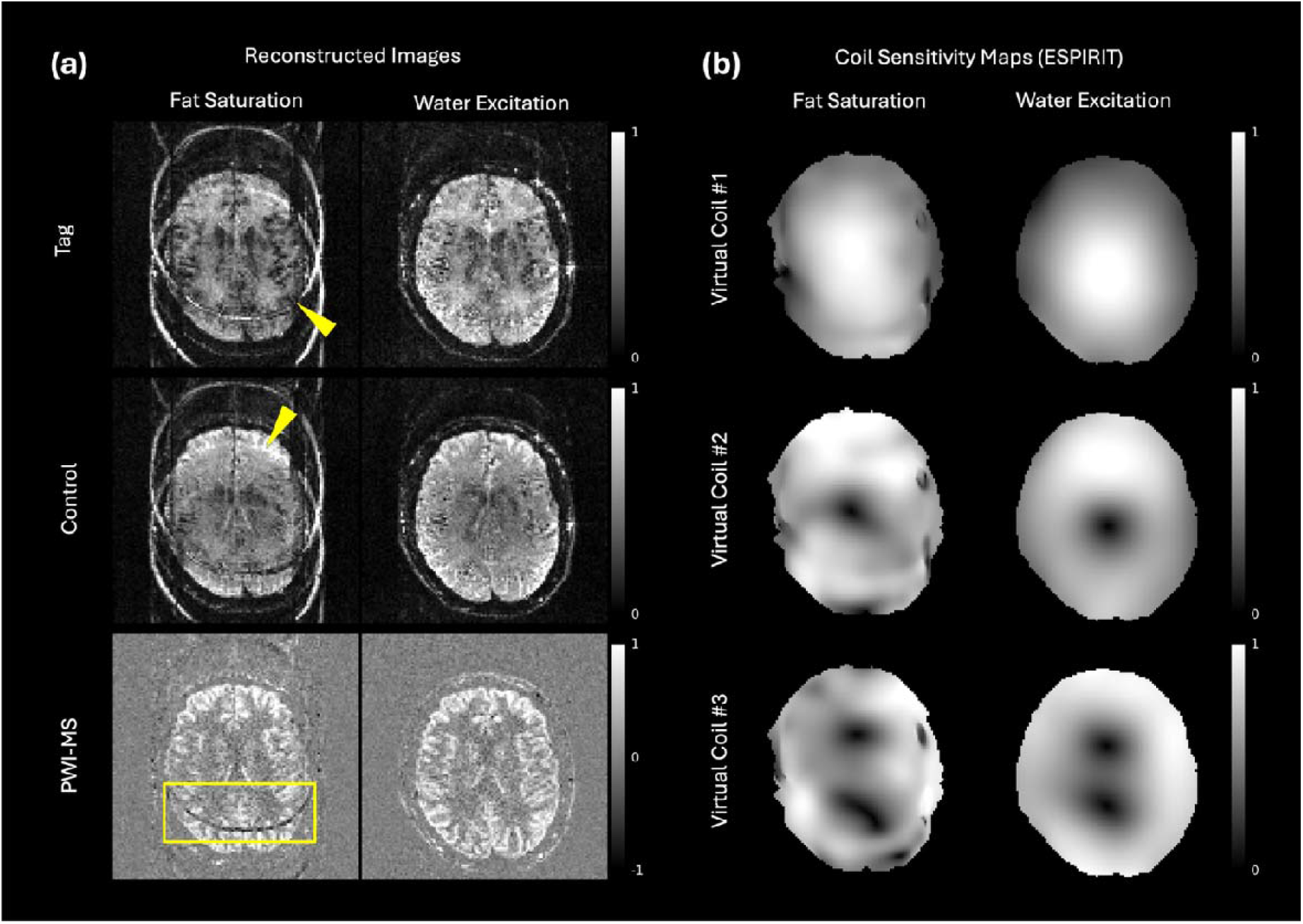
Effect of fat suppression method on reconstructed images and coil sensitivity maps. (a) Reconstructed tag, control, and perfusion-weighted images (magnitude subtraction) from a single subject using conventional fat saturation (left) and water excitation (right). Yellow arrows indicate lipid signal contamination in the tag and control images under fat saturation; the yellow box highlights the corresponding artifact in the perfusion image. (b) ESPIRiT-estimated coil sensitivity maps for three representative virtual coils under fat saturation (left) and water excitation (right). Residual lipid signal under fat saturation introduces structured errors in the estimated sensitivity maps, which propagate into the reconstructed images.

The residual lipid signal also affected coil sensitivity estimation. As shown in **Figure 3b**, ESPIRiT^44^-based sensitivity maps obtained under fat saturation exhibited spatial inconsistencies and structured distortions, whereas water excitation produced smooth and more physically plausible sensitivity profiles across all virtual coils. Because these sensitivity maps enter the reconstruction forward model, errors in their estimation propagate into every reconstructed image, making robust fat suppression a prerequisite for the strongly suppressed regime rather than a cosmetic improvement.

Additional binomial pulse comparisons supported the use of 1-2-1 water excitation as a balance between fat suppression and excitation duration (**Figure S2**).

### Subtraction Strategy

Figure 4 compares PWI obtained with magnitude subtraction (MS), complex subtraction (CS) under Strong BGS and a reference-superimposed approach followed by magnitude subtraction (RS). Under Strong BGS the tag and control images operate in a low-signal regime in which the residual longitudinal magnetization can approach zero or become negative. MS discards the sign of this magnetization, so where tag and control have opposite polarity, the subtraction is biased and fine perfusion structure is lost or distorted.

Both RS and CS recovered this structure, but by different routes. CS retains the phase and therefore the sign directly. RS instead adds the *M*_0_ signal to lift the tag and control images away from the null point before the magnitude is taken, so that the data phase is essentially aligned and the sign ambiguity does not arise. The two gave comparable perfusion image quality (**Figure 4**), but RS additionally suppressed background noise, most visibly outside the brain, because the superimposed *M*_*0*_ signal moves the measurement away from the low-signal regime in which magnitude data are noise-rectified. RS was therefore adopted for all subsequent analyses, with CS retained as a comparison condition (Supporting Information **Figure S5**).

**Figure 4.**
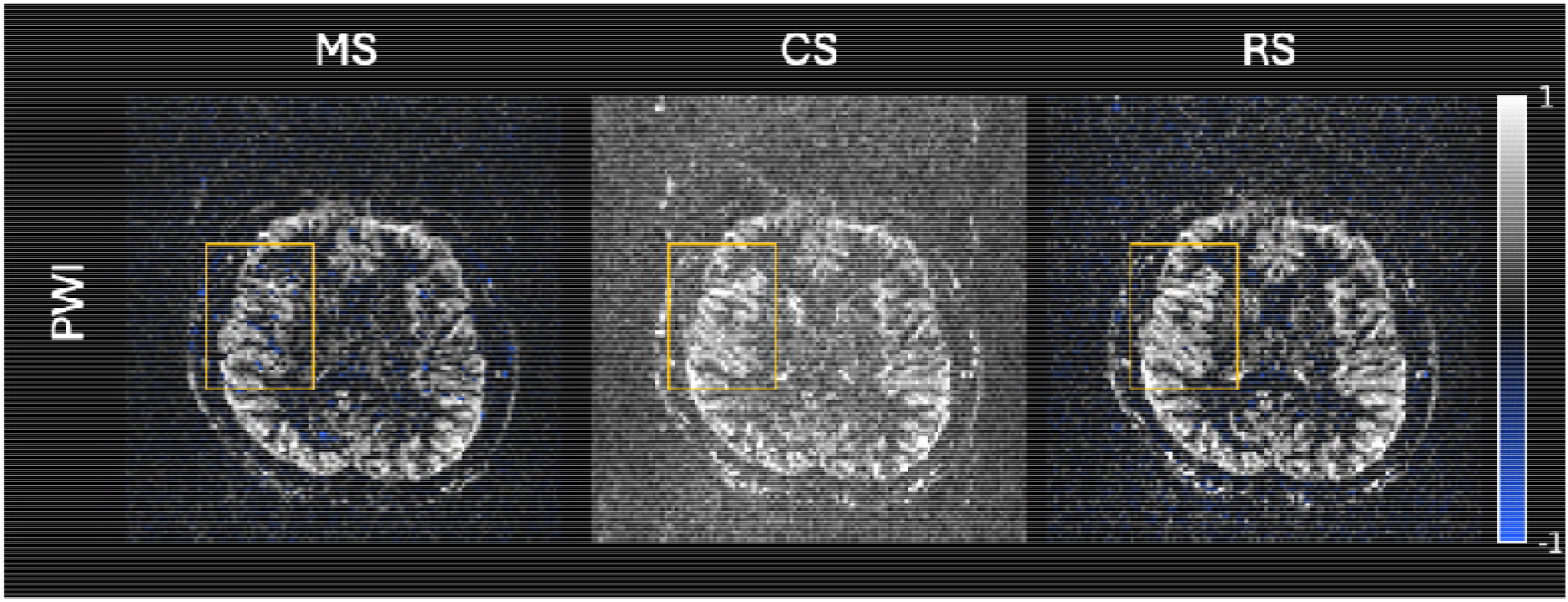
Effect of subtraction strategy on the perfusion-weighted image. Representative PWI from a single subject (motion-free data) obtained with three subtraction strategies: magnitude subtraction (MS), complex subtraction (CS), and reference-superimposed processing followed by magnitude subtraction (RS). Images were normalized using a shared positive reference defined by the 99.5th percentile of positive voxel values and displayed on a common scale from -1 to 1, with negative values encoded in blue and positive values in grayscale. Under Strong BGS the residual longitudinal magnetization can approach or cross zero, so MS exhibits sign-flip artifacts (orange box). CS restores the sign and RS lifts the signal away from the null point before the magnitude is taken, each recovering perfusion structure lost by MS. RS additionally suppresses background noise and was adopted for all subsequent analyses.

### Motion Correction

Figure 5 shows representative in vivo images from a single subject under Moderate BGS. Inter-shot motion introduced blurring and severe artifacts in the control and tag images, with more pronounced degradation in the PWI. The proposed alignedSENSE-PR reconstruction recovered sharp structural boundaries in the control and tag images and restored most of the perfusion signal, improving PWI quality over the motion-corrupted data (PSNR 21.3 to 23.3 dB, SSIM 0.297 to 0.517). Compared with the motion-free reference, the corrected images remained noisier but showed markedly improved structural definition relative to the motion-corrupted data.

Figure 6 presents the same comparison under Strong BGS. Despite the strongly reduced anatomical signal, alignedSENSE-PR recovered clear grey matter delineation and improved PWI quality (PSNR 24.3 to 25.3 dB, SSIM 0.474 to 0.604), demonstrating that accurate motion correction remains achievable in the low-background regime that is most challenging for conventional registration. Consistent with the BGS analysis above, the motion-corrupted Strong BGS data showed fewer tissue-related artifacts than Moderate BGS, reflecting the more complete suppression of static signal. Under complex subtraction, alignedSENSE-PR outperformed the inter-shot-only (alignedSENSE-IS) and mcFLIRT-based (alignedSENSE-MF) variants under both BGS conditions and the ablation is reported in Supporting Information **Figure S3**.

The estimated motion parameters in **Figure 7** clarify why the proposed method remains reliable under Strong BGS. Under Moderate BGS, all three variants produced broadly consistent rigid-body trajectories. Under Strong BGS, the large intensity difference between tag and control images destabilized the image-based registration step in alignedSENSE-MF, producing increased variability and local inconsistencies in the estimated parameters. alignedSENSE-PR instead produced smooth, consistent trajectories that agreed closely with those obtained under Moderate BGS, in line with the matched motion instructions used across the two acquisitions. Because alignedSENSE-PR estimates motion directly from the multicoil k-space data through the forward model rather than from image-domain tissue contrast, its motion estimates do not degrade when the background is suppressed.

**Figure 5.**
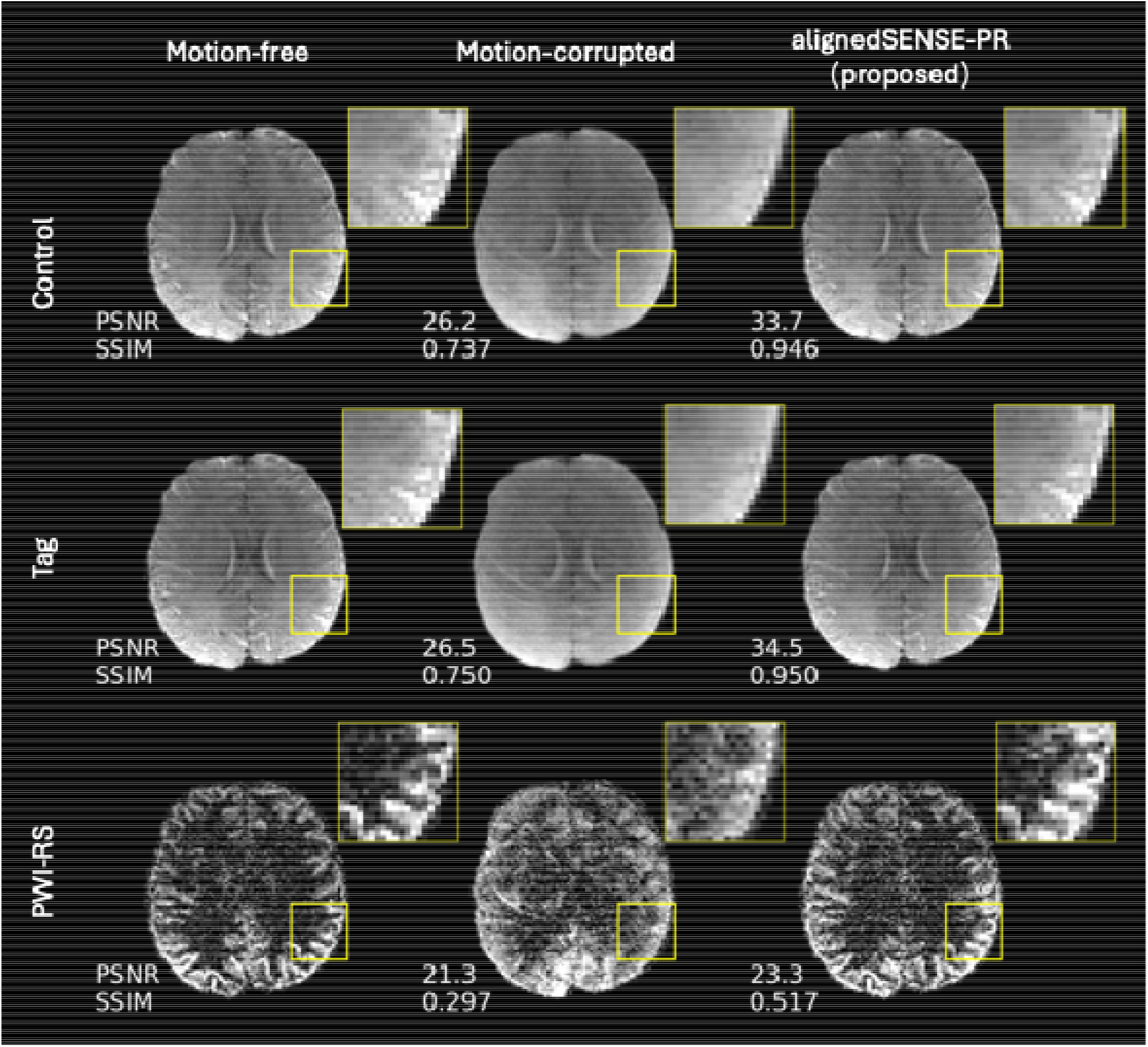
Motion correction results under Moderate BGS. Representative control, tag, and the perfusion-weighted images (PWI-RS) from a single subject under Moderate BGS, shown for three conditions: motion-free reference, motion-corrupted data, and alignedSENSE-PR. PSNR (dB) and SSIM values computed against the motion-free reference are annotated.

**Figure 6.**
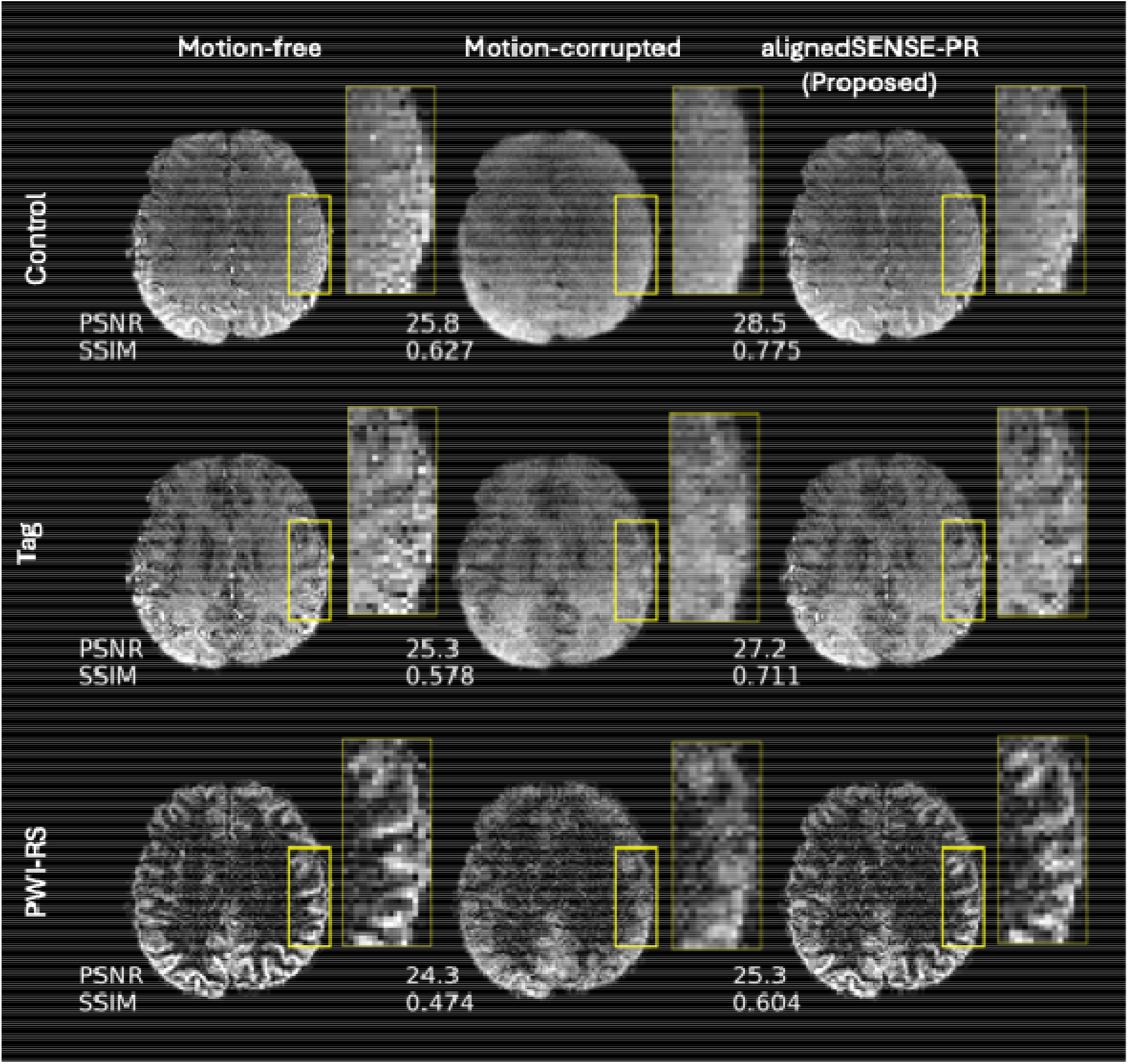
Motion correction results under Strong BGS. Representative control, tag, and perfusion-weighted image (PWI-RS) from the same subject under Strong BGS, shown for the motion-free reference, the motion-corrupted data and alignedSENSE-PR. PSNR (dB) and SSIM against the motiosn-free reference are annotated. Despite the strongly suppressed static-tissue contrast, alignedSENSE-PR recovers grey matter delineation and perfusion signal, with reduced artifacts in the PWI.

**Figure 7.**
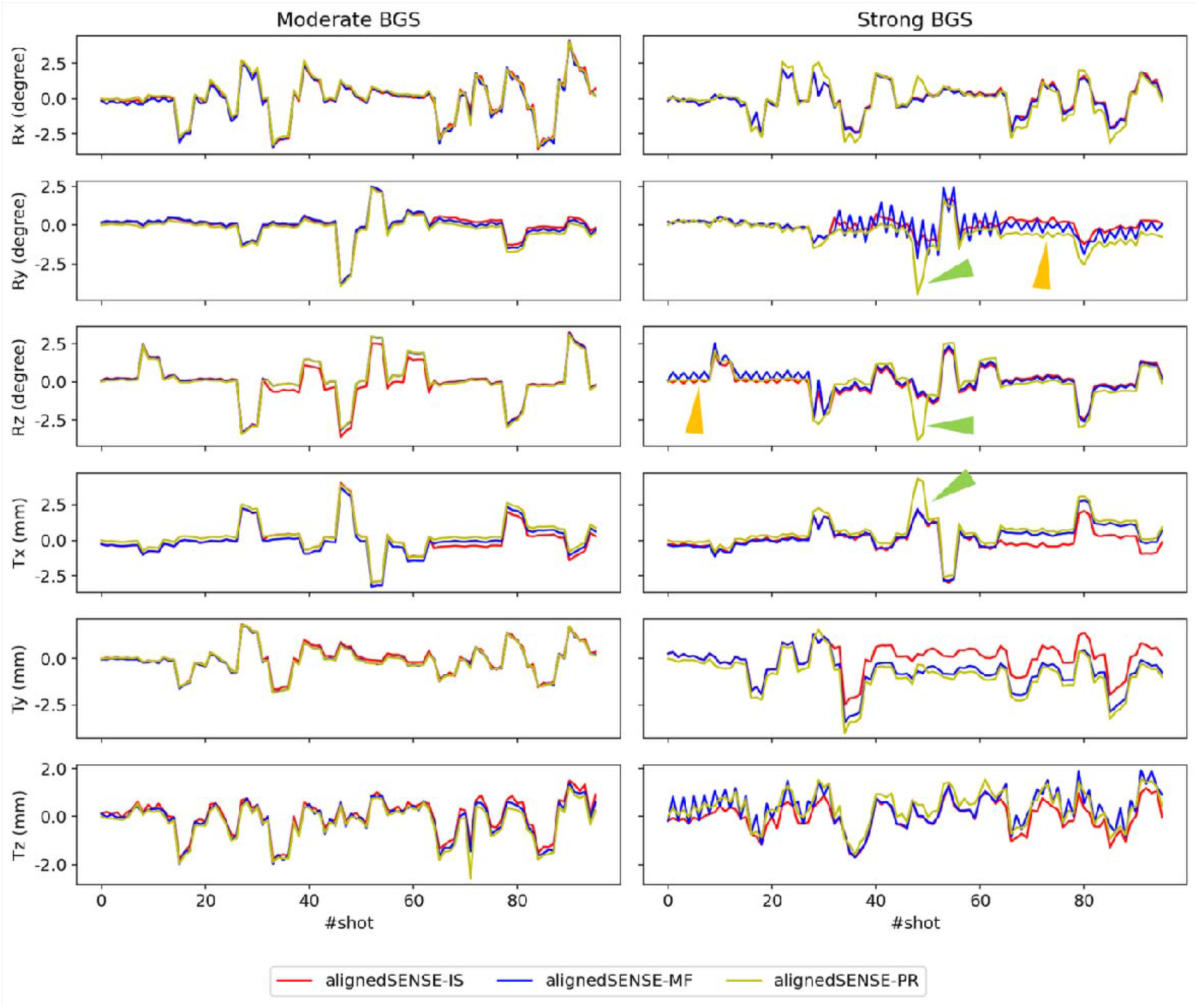
Estimated inter-shot rigid-body motion parameters. Six motion parameters (rotations Rx, Ry, Rz in degrees; translations Tx, Ty, Tz in mm) estimated across all k-space segments by three alignedSENSE variants (IS: red; MF: blue; PR: yellow), shown for Moderate BGS (left) and Strong BGS (right). Under Strong BGS, the large tag-control intensity difference destabilizes the image-based registration in alignedSENSE-MF, producing increased variability and local inconsistencies (orange arrows). In contrast, alignedSENSE-PR yields smooth trajectories that remain consistent with those estimated under Moderate BGS, including at several segments where it departs from both IS and MF (green arrows).

Figure 8 summarizes the perfusion image metrics across all eight subjects. alignedSENSE-PR significantly improved PSNR and SSIM relative to the motion-corrupted data under both Moderate and Strong BGS (paired-sample t-tests). Higher mean PSNR and SSIM were observed under Strong BGS than Moderate BGS across the cohort. The same motioncorrection benefit was observed when CS with LLR denoising was used in place of RS (Supporting Information **Figure S6**), indicating that the improvement from alignedSENSE-PR is independent of the subtraction strategy.

Figure 9 contrasts the two BGS schemes in the same subject: in the motion-free condition the two schemes were comparable, whereas under motion Moderate BGS showed greater loss of perfusion contrast and structural definition. After correction, alignedSENSE-PR recovered perfusion image quality in both conditions, with stronger recovery under Strong BGS, particularly in cortical regions.

## DISCUSSION

This work evaluated a set of sequence and reconstruction design choices for motion-robust high-resolution 3D-GRASE pCASL at 3T. We found that some of these choices interact with others, such as the use of strong background suppression, which yields cleaner perfusion measurement but is more challenging for conventional image-based motion correction. The reconstruction that estimates motion without relying on static-tissue contrast can use Strong BGS to its full advantage with water excitation and reference-superimposed processing for further reducing artifacts. The combination of Strong BGS, water excitation, reference-superimposed processing, and alignedSENSE-PR reconstruction provided robust performance in the presence of head motion.

**Figure 8.**
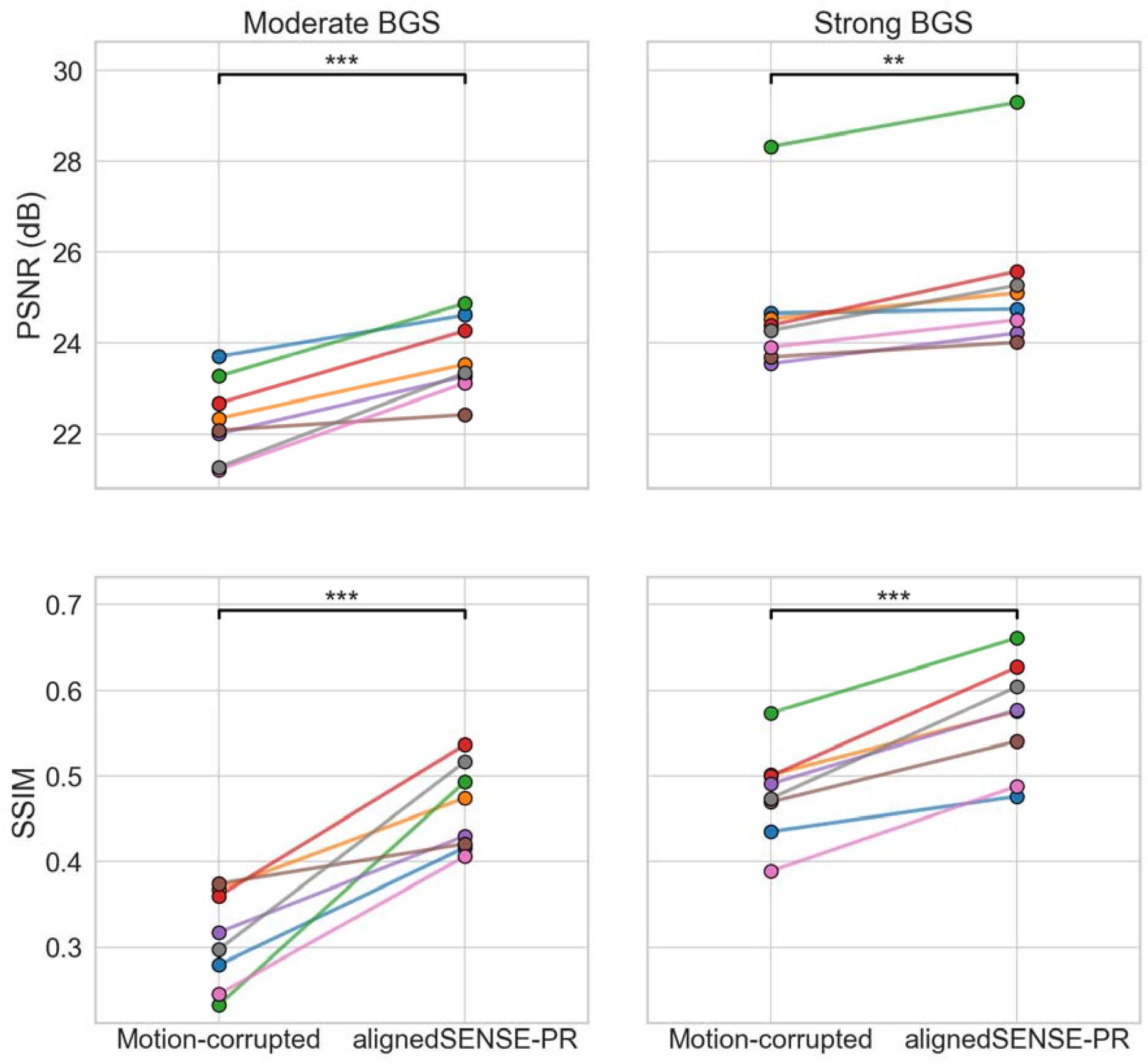
Quantitative motion correction across all subjects. PSNR (top) and SSIM (bottom) of the perfusion-weighted image (PWI-RS) computed against the motion-free reference for all eight subjects (individual-colored lines) under Moderate BGS (left) and Strong BGS (right), comparing the motion-corrupted data with alignedSENSE-PR. The motion-free reference and both conditions were processed with reference-superimposed (RS) correction. Statistical comparisons used paired-sample t-tests (*: p < 0.05, **: p < 0.01, ***: p < 0.001, ns: not significant).

**Figure 9.**
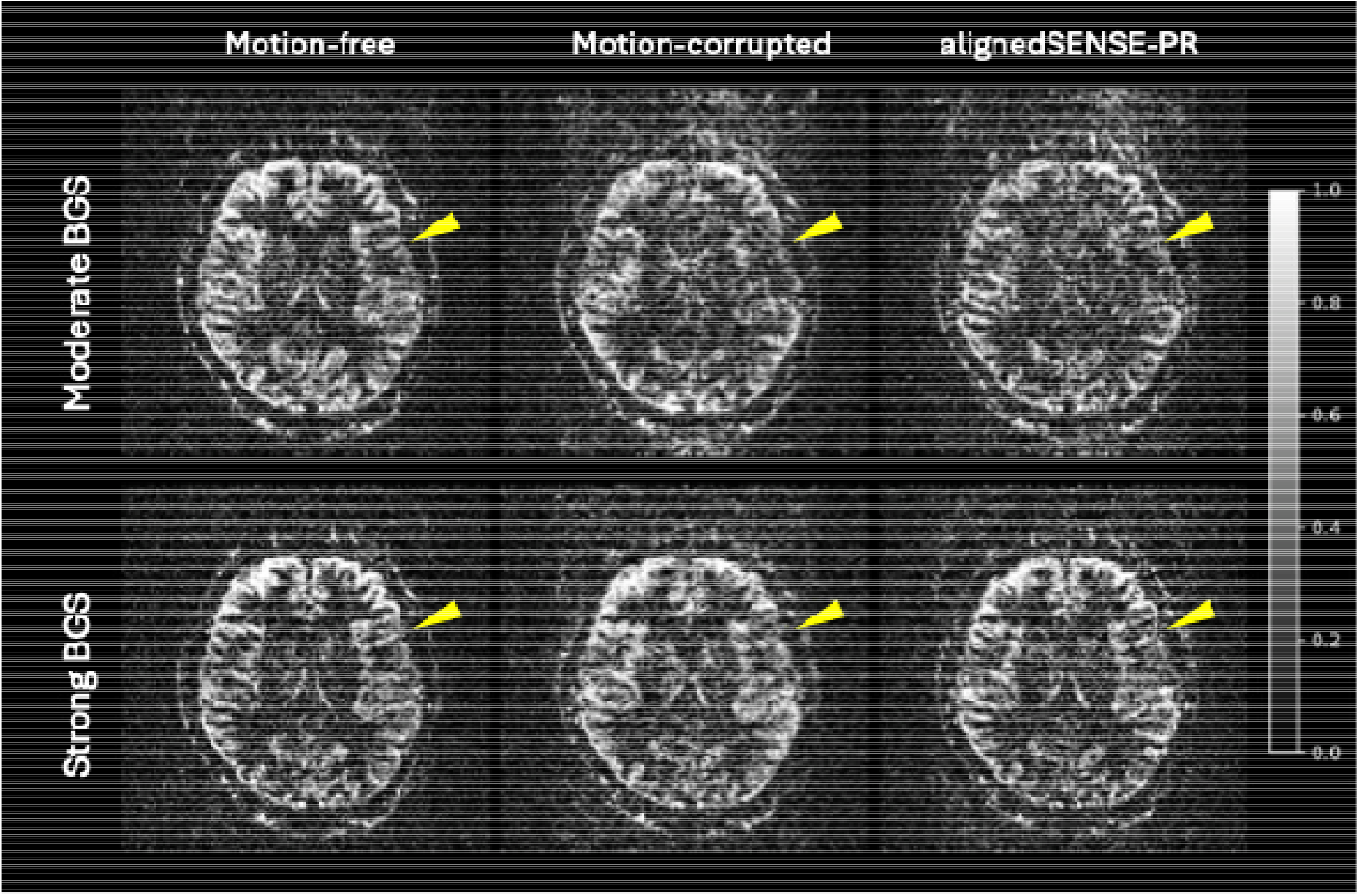
Perfusion-weighted image comparison across BGS conditions for a representative subject. Perfusion-weighted images (PWI-RS) from one subject under Moderate BGS (top row) and Strong BGS (bottom row), shown for three conditions: motion-free reference, motion-corrupted, and alignedSENSE-PR. Yellow arrows indicate a region of interest where motion artifacts and BGS-dependent differences are most apparent. Strong BGS additionally suppresses residual background signal outside the brain, reducing the artifacts visible in the surrounding region compared with Moderate BGS.

Motion-corrected reconstruction becomes increasingly important as spatial resolution increases, because higher-resolution segmented 3D-GRASE acquisitions require more shots and therefore longer acquisition times, increasing vulnerability to inter-shot motion. Each shot is too heavily undersampled to reconstruct the self-navigator for motion estimation^28^. alignedSENSE-PR addresses this by estimating inter-shot motion directly from the multi-coil k-space data through the reconstruction forward model, jointly with the image. Because the motion estimation is driven by consistency of the multi-coil data rather than by image-domain tissue contrast, it does not degrade when the static background is suppressed. And alignedSENSE-PR also simultaneously achieved the inter-volume correction by encouraging the minimum difference between tag and control images during the reconstruction. This allows accurate motion correction to persist into the Strong BGS regime, where the reduced anatomical signal and the large tag-control intensity difference make image-based registration unreliable.

With motion correction no longer dependent on the residual tissue contrast, the primary disadvantage of Strong BGS is eliminated. A principal benefit of the 4-inv BGS scheme is the more effective suppression of signals from all tissue types, particularly CSF, which is an important source of physiological fluctuation arising from pulsatility^32^. Such CSF-related signal cannot be fully removed by control-tag subtraction and can contaminate the perfusion signal through partial-volume effects. By suppressing static tissue signal more completely at excitation, Strong BGS also reduces the amount of background signal that can be converted into subtraction artifacts when motion occurs. This likely explains the reduced corruption observed in the motion-contaminated perfusion images and the improved recovery after reconstruction. Therefore, Strong BGS should be considered not as a harder regime, but as one that is intrinsically cleaner for the perfusion measurement while shifting the challenge onto the reconstruction that our proposed alignedSENSE-PR is designed to overcome.

Adopting strong suppression places specific demands on the rest of the pipeline. The first concerns fat. Once tissue is strongly suppressed, lipid becomes relatively more conspicuous and can emerge as a dominant structured contaminant in both the source images and the reconstructed perfusion images. This behavior is consistent with the simulations shown in **Figure 2**, where fat recovered substantially faster than the suppressed tissue compartments. Conventional fat saturation is vulnerable to failure because it relies on spectrally selective presaturation, which is sensitive to B0 and B1 inhomogeneity and frequency offsets arising from B0 drift across repetitions, all of which can produce spatially variable residual lipid signal. Water excitation is also sensitive to off-resonance, but in our implementation, it provided more stable fat suppression, likely because it is less sensitive to B1 variation and does not rely on the timing between fat saturation and excitation. This reduced structured chemically-shifted residual contamination in the strongly suppressed data. This advantage comes with known trade-offs, including sensitivity to off-resonance and a longer TE, particularly for longer binomial schemes. Nevertheless, within this trade-off space, the 1-2-1 water-excitation scheme provided the best overall balance between robustness and image quality in the present implementation.

The second demand concerns the subtraction. Under Strong BGS the control and tag image intensities approach zero and can reverse in sign. Magnitude subtraction discards this sign, leading to biased subtraction and structural distortion. Reference-superimposed processing resolves this by adding the *M*_*0*_ signal to lift the tag and control images away from the null point before the magnitude is taken, additionally suppressing background noise. Complex subtraction resolves the same ambiguity by retaining the phase and resulted in comparable image quality without requiring an *M*_0_ reference. Reference-superimposed processing is particularly well suited to this framework because alignedSENSE-PR brings the tag and control data into the *M*_*0*_ frame as part of the reconstruction itself, rather than requiring a separate registration step. Otherwise, any residual misalignment between *M*_*0*_ and the tag/control data would introduce extra error. Reference-superimposed processing therefore places a stricter alignment requirement on the pipeline than complex subtraction.

The optional locally low-rank constraint occupies a different role from the choices discussed above. Because the perfusion component is highly redundant across repeats, a low-rank reconstruction provides additional noise suppression under Strong BGS, where perfusion SNR is lowest, and the motion-correction benefit of alignedSENSE-PR is preserved under this back-end (Supporting Information **Figure S6**). It was deliberately excluded from the main analysis so that the reported image quality reflects the acquisition and reconstruction choices themselves rather than a subsequent denoising step, but it remains available where SNR, rather than fidelity to the underlying perfusion distribution, is the limiting factor.

Across subjects, Moderate BGS tended to produce more severe motion-related artifacts and less complete recovery of the perfusion signal after correction than Strong BGS. This indicates that Strong BGS is intrinsically more robust to motion contamination, despite placing greater demands on motion estimation. In other words, Strong BGS leaves less residual background from static tissue and CSF to be transformed into subtraction artifacts, so once motion is handled the perfusion signal is recovered more fully than under a moderate regime. Together these observations support Strong BGS, combined with alignedSENSE-PR, as the preferred strategy rather than the conservative suppression levels typically chosen to protect retrospective registration.

Several limitations should be acknowledged. First, the study included only eight healthy volunteers and employed instructed motion rather than naturally occurring motion. Although the cued-motion paradigm provided a controlled way to test robustness, motion amplitude was not strictly standardized across subjects, and the observed motion may not fully represent involuntary or pathological motion encountered in clinical practice. Second, the image quality was assessed against an assumed noise- and motion-free reference. Third, the current study focused on technical optimization and did not directly assess how the improved highresolution perfusion images translate into specific clinical applications.

Future work should therefore evaluate this framework in patient cohorts and determine whether the observed gains in image robustness translate into more accurate quantitative cerebral blood flow estimation. It will also be important to investigate whether the improved spatial fidelity of these high-resolution perfusion images enables clinically meaningful benefits, for example in resolving cortical or periventricular perfusion abnormalities that are otherwise obscured by partial-volume effects.

## CONCLUSIONS

This work demonstrated that combining strong background suppression with the proposed alignedSENSE-PR reconstruction can achieve motion-robust high-resolution 3D-GRASE pCASL at 3T. Strong BGS leaves less residual static-tissue and CSF signal, reducing contamination of the perfusion image, while alignedSENSE-PR estimates and corrects motion reliably even when static background is strongly suppressed. Together with water excitation and reference-superimposed processing, this combination enabled robust 2.0 mm isotropic perfusion imaging within a clinically acceptable scan time.

## Supporting information

Support information

## Data Availability

The data supporting the findings of this study are not publicly available due to participant privacy and ethical restrictions.

## Acknowledgments

This work conducted in the Oxford Centre for Integrative Neuroimaging was supported by core funding from the Wellcome Trust (203139/Z/16/Z and 203139/A/16/Z) with additional support from the NIHR Oxford Biomedical Research Centre (NIHR203311) and the Oxford Health Biomedical Research Centre (NIHR203316). M.H. is supported by the Jardine Foundation. M.C. is supported by the Canada Research Chairs Program. P.J. is supported by the Vivensa Foundation and the NIHR Oxford Biomedical Research Centre (NIHR203311). T.O. was supported by a Sir Henry Dale Fellowship jointly funded by the Wellcome Trust and the Royal Society (220204/Z/20/Z) and the Podium Institute for Sports Medicine and Technology, University of Oxford. For the purpose of open access, the author has applied a CC BY public copyright license to any Author Accepted Manuscript version arising from this submission.

## Conflict of Interest Statement

PJ is the Editor-in-Chief of Magnetic Resonance in Medicine. In line with COPE guidelines, he recused himself from all involvement in the review process of this paper, which was handled by an associate editor. He and the other authors had no access to the identities of the reviewers.

## Notes

### Competing Interest Statement

The authors have declared no competing interest.

### Author Declarations

The Clinical Trials and Research Governance Department of the University of Oxford gave ethical approval for the technical development protocol used in this work.

