## Supplementary material for "Motion-robust high-resolution 3D-GRASE pseudo-continuous ASL perfusion imaging at 3T": Support information

### Supporting Information

#### Text S1. LLR denoising stage

An additional option within the second stage perfusion image reconstruction is to further denoise the perfusion image using an additional regularization constraint. The inter-shot and inter-volume rigid-body transforms estimated in the first stage were held fixed, and each repeat was given its own $x_{r}$ and ${\Delta x}_{r}$ so that redundancy of the perfusion component across repeats could be exploited by a locally low-rank (LLR) constraint for denoising under the low SNR conditions of Strong BGS:

$$\min_{x_{r},{\Delta x}_{r}} \sum_{r} \sum_{i} \left( \left\| M_{i}FST_{t,r,i}(x_{r}-{\Delta x}_{r})-y_{t,r} \right\|_{2}^{2}+\left\| M_{i}FST_{c,r,i}(x_{r}+{\Delta x}_{r})-y_{c,r} \right\|_{2}^{2} \right)+ \lambda\sum_{p} \left\| C_{p}D \right\|_{*}$$

where $C_{p}$ extracts a $4\times4\times4$ patch (stride $2\times2\times2$) and $D$ is the $64\times$ 3 Casorati matrix stacking the patch voxels of $\left\{ \Delta x_{r} \right\}, r=\{1, 2, 3\}$, $\left\| \cdot\right\|_{*}$ is the nuclear norm. Coil sensitivities were reused from the first stage. The problem was solved by variable splitting: each outer iteration alternated a data-consistency image update solved by conjugate-gradient algorithm with an LLR proximal step applied as patch-wise singular-value soft-thresholding, for up to 100 outer iterations with tolerance ${10}^{-6}.$

The LLR weight $\lambda=0.5$for Strong BGS data and $\lambda=0.18$for Moderate BGS, which was selected empirically to balance denoising against preservation of perfusion structure (see **Figure S4**). A stronger weight was used for Strong BGS, where the lower perfusion SNR requires more aggressive denoising, than for Moderate BGS, where more residual background signal is retained and a smaller weight preserves fine perfusion detail. The selected values gave stable image quality for both the motion-free and motion-corrected data.

The two-stage design deliberately separates motion estimation from denoising. In the first stage, sharing one perfusion image across repeats provides the global consistency needed for robust motion estimation under a simple, well-conditioned L2 model, avoiding the nonconvex interaction between a low-rank prior and the motion parameters. In the second stage, reconstructing per-repeat images exposes the inter-volume redundancy, allowing the low-rank constraint to achieve stronger denoising than L2 regularization.

**The role of the perfusion regularization is shown in Figure S1, evaluated at the motion-estimation stage with L2 penalty on the perfusion image. Reducing the regularization weight to** $\alpha= 0.1$ **introduced fluctuations in the estimated motion parameters (Figure S1b) and amplified noise in the PWI, lowering SSIM and PSNR. Increasing** $\alpha$ **to 5 over-regularized the perfusion estimate and suppressed perfusion signal in the difference map. At** $\alpha= 1$ **the motion trajectories were stable and the residual difference map was unstructured, indicating well-preserved perfusion contrast. The destabilization of motion estimation at low** $\alpha$ **confirms that the perfusion regularization does more than denoise. By penalizing the spurious static-tissue subtraction that arises under residual misalignment, it stabilizes motion estimation precisely where the large tag-control contrast change of Strong BGS would otherwise corrupt it.** $\alpha= 1$ **was therefore used for all motion estimation.**

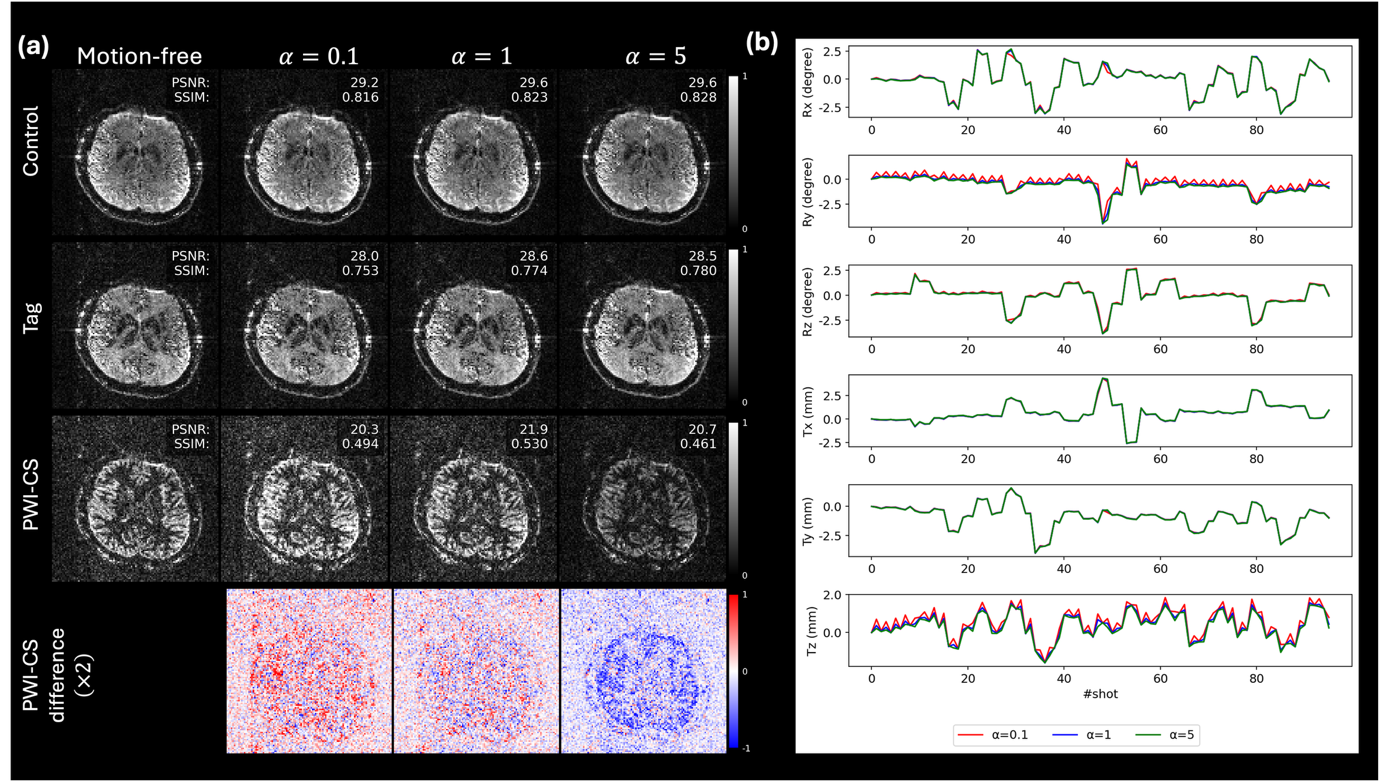

**Figure S1 Effect of the perfusion regularization weight α on motion estimation.** (a) Representative control, tag, PWI, and perfusion difference (×2 intensity, blue–red colormap) images for α = 0.1, 1, and 5, with the motion-free reference shown for comparison. All images are from the stage-1 reconstruction (L2 penalty on the perfusion image, without LLR denoising), isolating the effect of α on motion estimation. PSNR (dB) and SSIM against the motion-free reference are annotated. The unstructured difference map at α = 1 indicates well-preserved perfusion signal, in contrast to noise amplification at α = 0.1 and signal suppression at α = 5. **(b)** Estimated inter-shot motion parameters (rotations Rx, Ry, Rz; translations Tx, Ty, Tz) for the three α values, demonstrating increased estimation variability at α = 0.1 and stable estimates at α = 1 and 5.

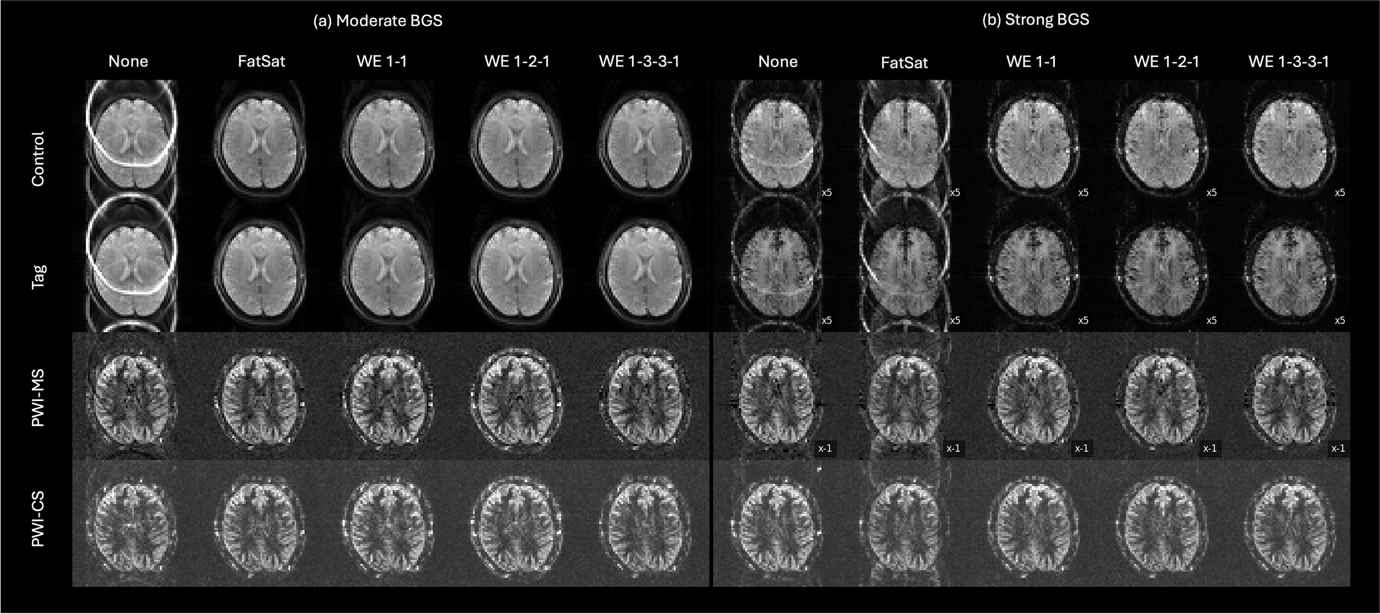

**Figure S2 Comparison of fat suppression strategies at reduced spatial resolution under Moderate and Strong BGS**. Representative control, tag, and perfusion-weighted images from a single subject acquired at reduced resolution (3.6 mm iso) under (a) Moderate BGS and (b) Strong BGS. Within each panel, columns correspond to five fat suppression strategies: no fat suppression (None), conventional fat saturation (FatSat), and three binomial water excitation schemes (WE 1-1, WE 1-2-1, WE 1-3-3-1). Rows show control, tag, perfusion (magnitude subtraction), and perfusion (complex subtraction) images. Lipid signal contamination visible under FatSat is progressively reduced with higher-order water excitation pulses. Higher-order pulses improved fat suppression but increased excitation duration and TE, whereas the WE 1-2-1 scheme provided the selected balance between suppression and readout efficiency.

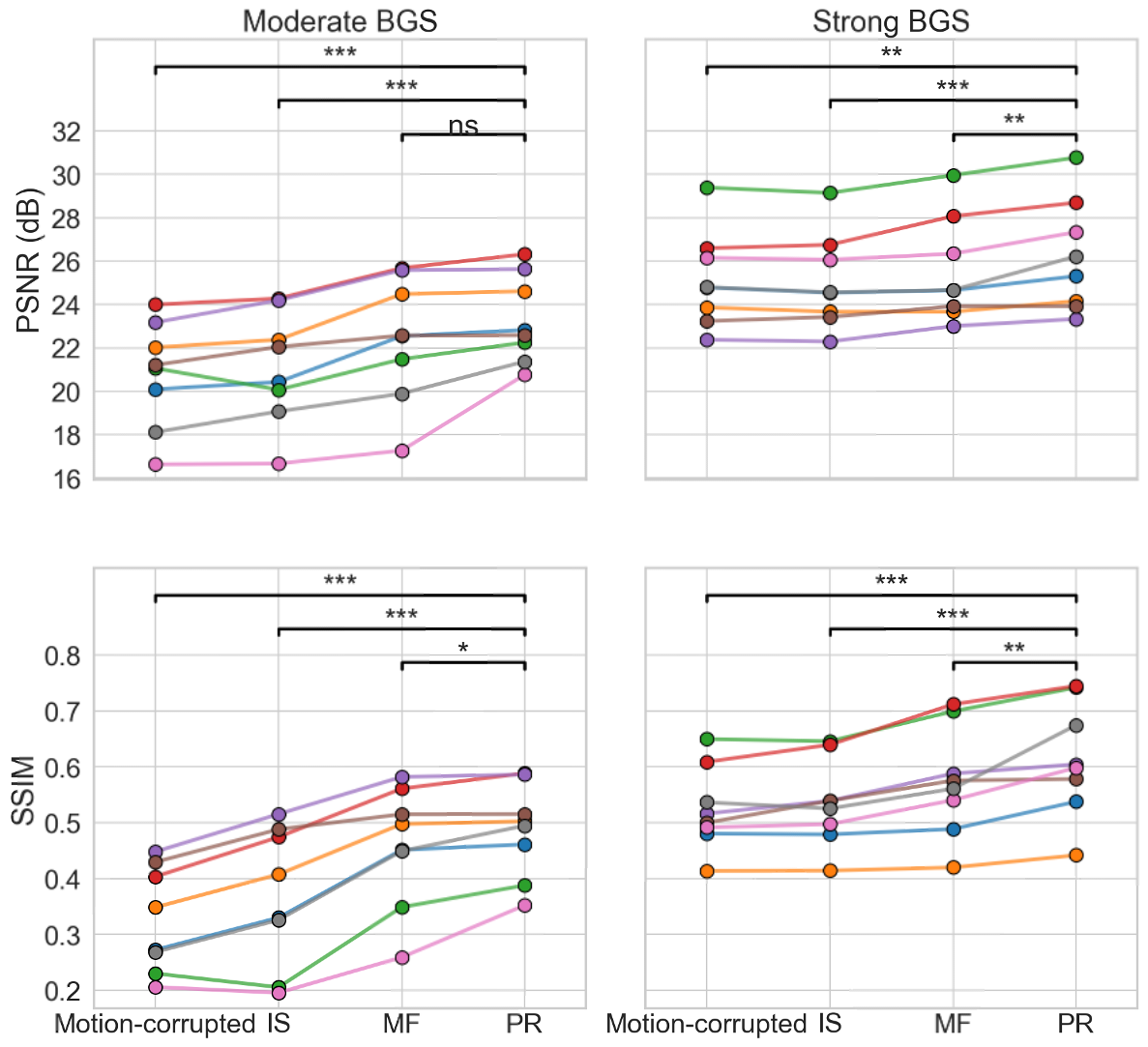

**Figure S3 Quantitative motion correction results across all subjects.** PSNR (top) and SSIM (bottom) of the perfusion-weighted image computed against the motion-free reference for all eight subjects (individual-colored lines) under Moderate BGS (left) and Strong BGS (right), across four reconstruction conditions: motion-corrupted, alignedSENSE-IS, alignedSENSE-MF, and alignedSENSE-PR. Metrics were computed on the complex-subtraction PWI with LLR denoising, so the absolute values are not directly comparable with Figure 8. Reference-superimposed processing is compatible only with alignedSENSE-PR, as it requires k-space alignment to the $M_{0}$ reference that the inter-shot-only and mcFLIRT variants cannot provide; complex subtraction, which applies to all variants, was therefore used for this ablation. Statistical comparisons were performed using paired-sample t-tests (*: p < 0.05, **: p < 0.01, ***: p < 0.001, ns: not significant).

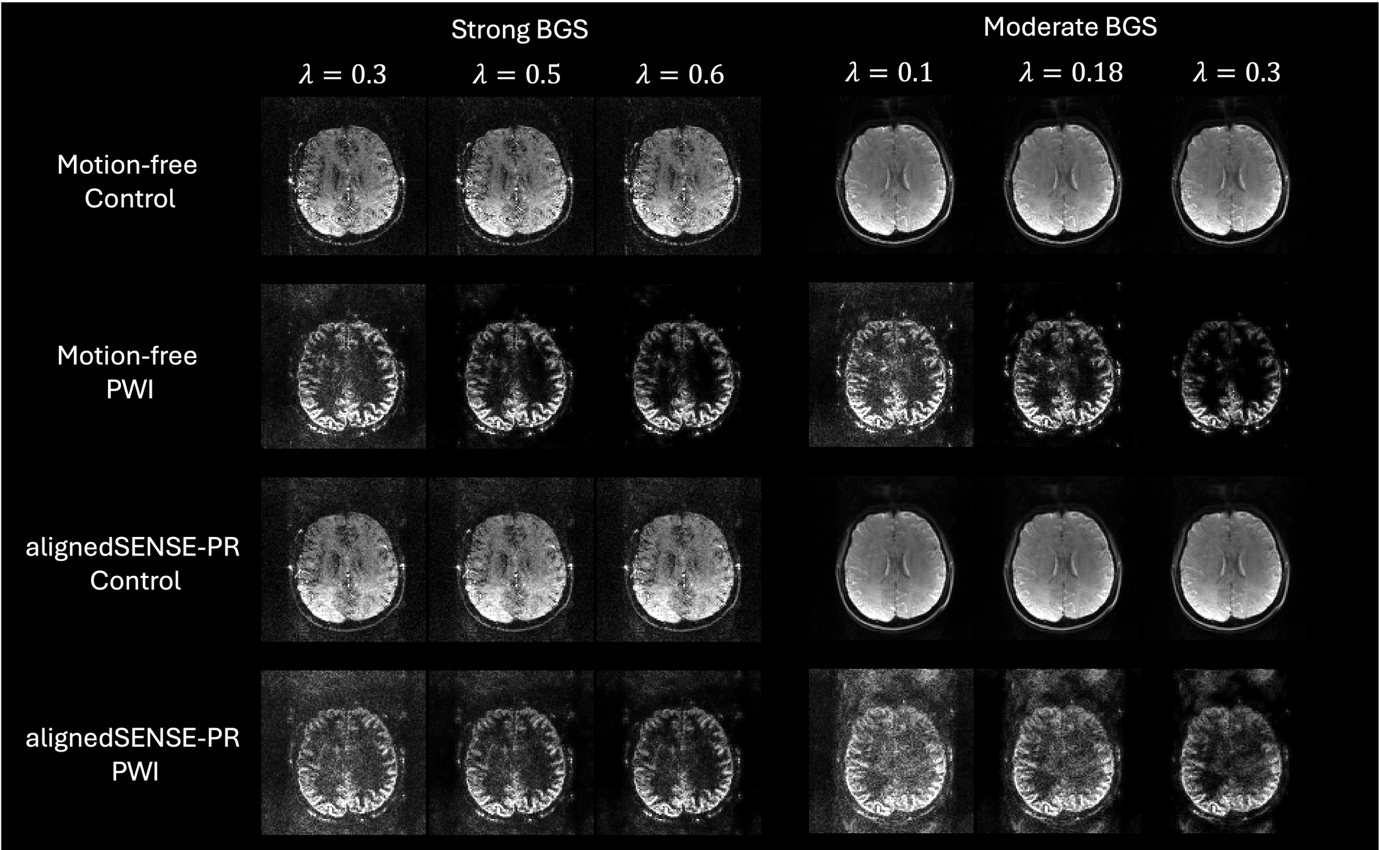

**Figure S4 Effect of the optional LLR denoising weight λ on reconstructed image quality.** Representative control and perfusion-weighted images (PWI) from a single subject reconstructed with a range of LLR weights, under Strong BGS (left, $\lambda= 0.3, 0.5, 0.6$) and Moderate BGS (right, $\lambda= 0.1, 0.18, 0.3$). The top two rows show the motion-free reference data and the bottom two rows the motion-corrected (alignedSENSE-PR) data. Increasing $\lambda$ progressively suppresses noise but oversmooths fine perfusion structure at the highest values. When LLR denoising is applied, $\lambda= 0.5$ for Strong BGS and $\lambda= 0.18$ for Moderate BGS balance denoising against preservation of perfusion detail.

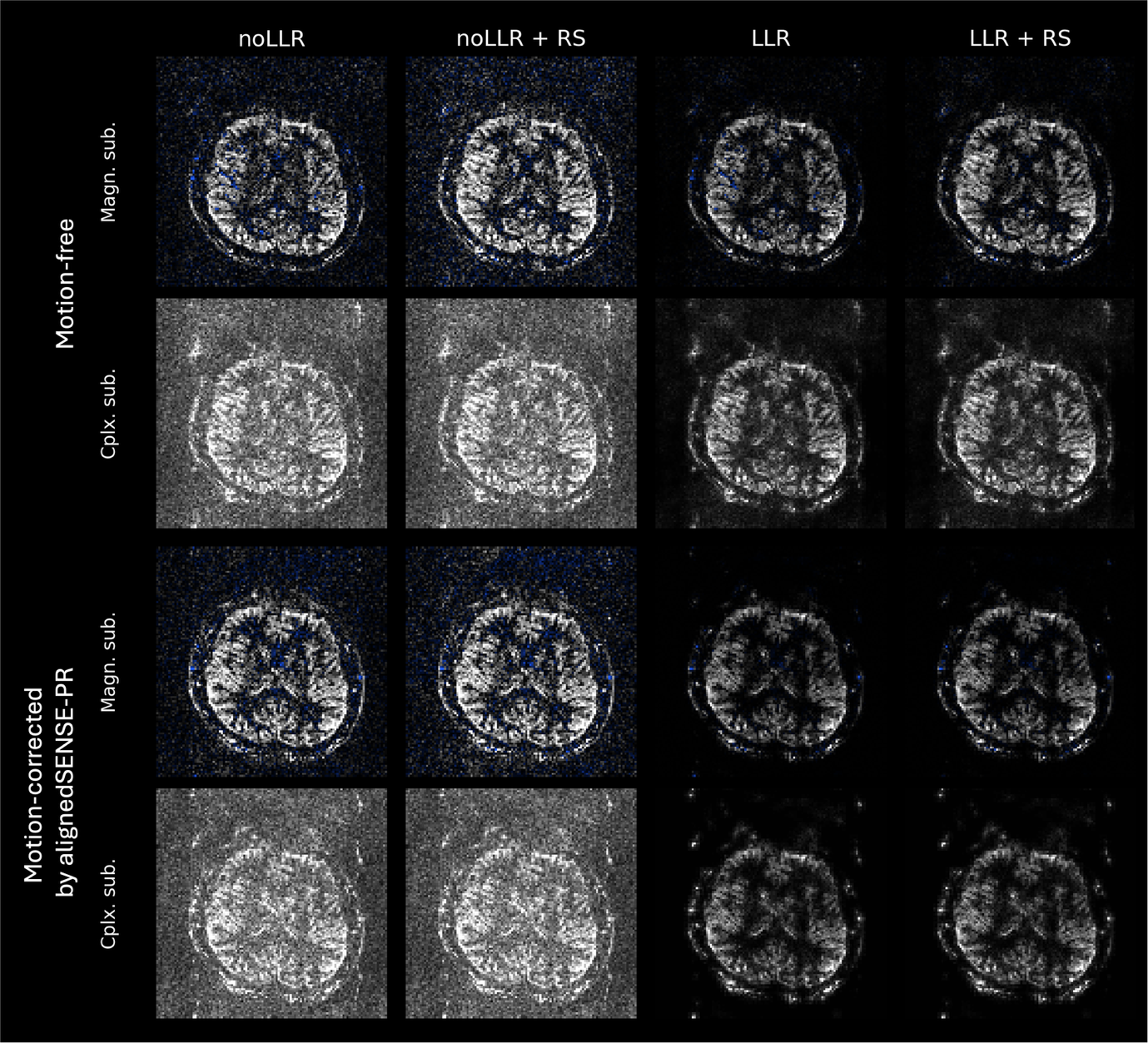

**Figure S5 Compatibility of reference-superimposed processing with LLR denoising under Strong BGS.** Representative perfusion-weighted images (PWI) from a single subject under Strong BGS, shown for motion-free data (top two rows) and motion-corrected data reconstructed with alignedSENSE-PR (bottom two rows). Within each, rows correspond to magnitude subtraction (MS) and complex subtraction (CS), and columns to four reconstruction back-ends: without LLR denoising (noLLR), reference-superimposed processing without LLR (noLLR with RS), LLR denoising (LLR), and reference-superimposed processing with LLR (LLR with RS). RS suppresses background noise on its own, most visibly for magnitude subtraction, which is why it is used without LLR in the main text. RS remains compatible with LLR denoising when stronger suppression is desired.

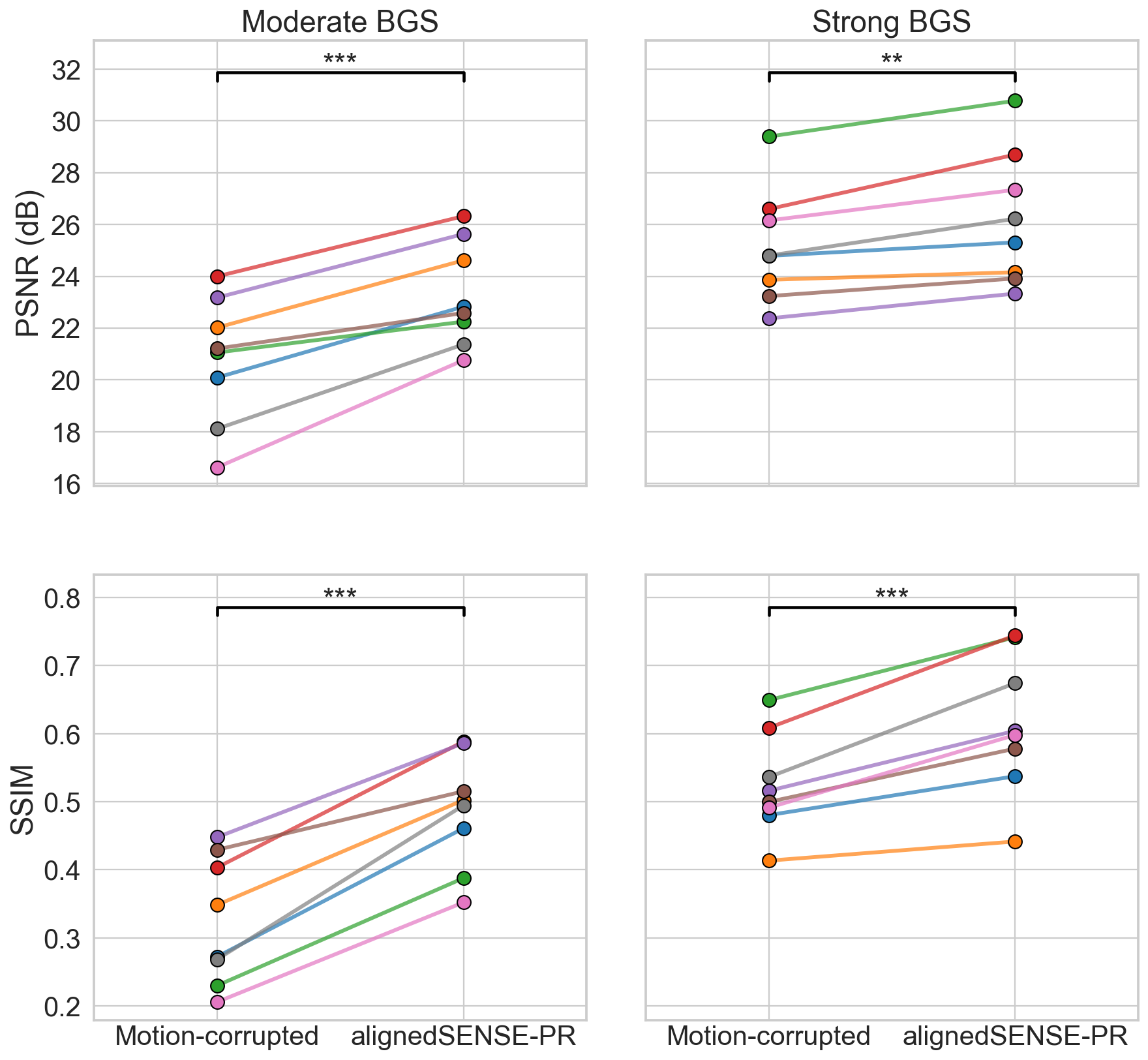

**Figure S6 Motion correction under complex subtraction with LLR denoising.** PSNR (top) and SSIM (bottom) of the PWI computed against the motion-free reference for all eight subjects (individual-colored lines) under Moderate BGS (left) and Strong BGS (right), comparing the motion-corrupted data with alignedSENSE-PR. Statistical comparisons used paired-sample t-tests (*: p < 0.05, **: p < 0.01, ***: p < 0.001, ns: not significant). alignedSENSE-PR improves both metrics under this back-end as well, showing that the motion-correction benefit does not depend on the choice of subtraction strategy.
